# Evaluation of the diagnostic value of YiDiXie™-SS, YiDiXie™-HS and YiDiXie™-D in pancreatic cancer

**DOI:** 10.1101/2024.07.30.24311246

**Authors:** Xutai Li, Pengwu Zhang, Hui Zhang, Chen Sun, Zhenjian Ge, Wenkang Chen, Yingqi Li, Shengjie Lin, Wuping Wang, Siwei Chen, Yutong Wu, Huimei Zhou, Wei Li, Fei Feng, Zewei Lin, Yongqing Lai

## Abstract

**Background:** Pancreatic cancer is a serious threat to human health. Enhanced CT is widely used in the diagnosis of pancreatic cancer. However, false-positive results on enhanced CT can lead to misdiagnosis and incorrect surgery or treatment, while false-negative results on enhanced CT can lead to missed diagnosis and delayed treatment. There is an urgent need to find convenient, cost-effective and non-invasive diagnostic methods to reduce the false-positive and false-negative rates of enhanced CT in pancreatic tumors. The aim of this study was to evaluate the diagnostic value of YiDiXie™-SS, YiDiXie™-HS and YiDiXie™-D in pancreatic cancer.

**Patients and methods:** 62 subjects (malignant group, n=37; benign group, n=25) were finally included in this study. Remaining serum samples from the subjects were collected and tested by applying the YiDiXie™ all-cancer detection kit to evaluate the sensitivity and specificity of YiDiXie™-SS, YiDiXie™-HS and YiDiXie™-D, respectively.

**Results:** The sensitivity of YiDiXie™ SS was 100% (95% CI: 90.6% - 100%) and its specificity was 68.0% (95% CI: 48.4% - 82.8%). This means that YiDiXie™ SS has extremely high sensitivity and high specificity in pancreatic tumors.YiDiXie™-HS has a sensitivity of 94.6% (95% CI: 82.3% - 99.0%) and a specificity of 84.0% (95% CI: 65.3% - 93.6%). This means that YiDiXie™-HS has high sensitivity and specificity in pancreatic tumors.YiDiXie™-D has a sensitivity of 73.0% (95% CI: 57.0% - 84.6%) and its specificity is 92.0% (95% CI: 75.0% - 98.6%). This means that YiDiXie™-D has high sensitivity and very high specificity in pancreatic tumors.YiDiXie™ SS had a sensitivity of 100% (95% CI: 88.6% - 100%) and a specificity of 66.7% (95% CI: 30.0% - 94.1%) in patients with positive enhanced CT. This means that the application of YiDiXie™ SS reduces the false-positive rate of enhanced CT by 66.7% (95% CI: 30.0% - 94.1%) with essentially no increase in malignancy leakage. YiDiXie™-HS had a sensitivity of 85.7% (95% CI: 48.7% - 99.3%) and a specificity of 84.2% (95% CI: 62.4% - 92.5%) in enhanced CT-negative patients. This means that YiDiXie™-HS reduces the false-negative enhancement CT rate by 84.2% (95% CI: 62.4% - 92.5%). YiDiXie™-D has a sensitivity of 73.3%(95% CI: 55.6% - 85.8%) and a specificity of 83.3%(95% CI: 43.6% - 99.1%) in patients with positive enhancement CT. This means that YiDiXie™-D reduces the false positive rate of enhanced CT by 83.3%(95% CI: 43.6% - 99.1%). YiDiXie™-D had a sensitivity of 71.4% (95% CI: 35.9% - 94.9%) and a specificity of 94.7% (95% CI: 75.4% - 99.7%) in patients with negative enhanced CT. This means that YiDiXie™-D reduces the false-negative rate of enhanced CT by 71.4% (95% CI: 35.9% - 94.9%) while maintaining high specificity.

**Conclusion:** YiDiXie ™ -SS has very high sensitivity and high specificity in pancreatic tumors.YiDiXie ™ -HS has high sensitivity and high specificity in pancreatic tumors.YiDiXie™-D has high sensitivity and very high specificity in pancreatic tumors.YiDiXie™ SS significantly reduces pancreatic-enhanced CT with essentially no increase in delayed treatment of malignant tumors. The YiDiXie™-HS significantly reduces the false negative rate of pancreatic enhanced CT. the YiDiXie™ -D significantly reduces the false positive rate of pancreatic enhanced CT or significantly reduces the false negative rate of pancreatic enhanced CT while maintaining a high level of specificity. the YiDiXie™ test is of great diagnostic value in pancreatic cancer, and it is expected to solve the problem of “ too high a false positive rate” and “ too high a false negative rate” of pancreatic enhanced CT.

**Clinical trial number:** ChiCTR2200066840.

## INTRODUCTION

Among the most prevalent malignant tumors is pancreatic cancer. According to recent data, 510,000 new instances of pancreatic cancer and 460,000 new deaths from the disease are expected worldwide in 2022^1^; the incidence and mortality of pancreatic cancer have increased by 2.9% and 0.2%, respectively, in 2022 compared to 2020^1,2^. Since there is no early warning sign of pancreatic cancer, many patients are diagnosed with advanced disease, which contributes to a high death rate even though total surgical resection is the only treatment that can cure the disease. A extremely low rate of surgical resectability at the time of diagnosis is also linked to this^3-5^. More than 80% of patients with pancreatic cancer are diagnosed with locally advanced or metastatic disease and are lost to surgery, with less than 20% surviving for 5 years, and the median survival for patients with advanced and metastatic disease is less than one year with treatment^6^. Thus, pancreatic cancer is a serious threat to human health.

Enhanced CT is widely used in the diagnosis of pancreatic tumors. On the one hand, enhanced CT can produce a large number of false-positive results. The false-positive rate of enhanced CT for the diagnosis of recurrent pancreatic cancer is about 24%-33%^7-10^. With a positive enhanced CT, patients usually undergo radical resection^11-12^. A false-positive result on enhanced CT means that a benign disease is misdiagnosed as a malignant tumor, and the patient will have to bear the undesirable consequences of unnecessary mental anguish, costly surgeries and investigations, surgical trauma, organ removal, and loss of function. Therefore, there is an urgent need to find a convenient, cost-effective and noninvasive diagnostic method to reduce the rate of false-positive pancreatic enhanced CT.

On the other hand, enhanced CT can produce a large number of false-negative results. The negative predictive value of enhanced CT for the diagnosis of pancreatic cancer is only 16.7%, and its false-negative rate is 26.3%^13^. In diagnosing recurrent pancreatic cancer tumors, the negative predictive value of enhanced CT is only 30-50%^7,9,10^, and its false-negative rate is about 25-27%^9,10^. When enhancement CT is negative, patients are usually taken for observation and regular follow-up^12^. A false-negative result on enhanced CT implies misdiagnosis of a malignant tumor as a benign disease, which will likely lead to delayed treatment, progression of the malignant tumor, and possibly even development of an advanced stage. Patients will thus have to bear the adverse consequences of poor prognosis, high treatment costs, poor quality of life, and short survival. Therefore, there is an urgent need to find a convenient, economical and noninvasive diagnostic method to reduce the false-negative rate of pancreatic enhancement CT.

In addition, there are some special patients who require extra caution in choosing whether to operate, e.g., smaller tumors, patients in poor general condition, etc. The risk of wrong surgery in these special patients is much higher than the risk of missed diagnosis. And the false-positive result of enhanced CT means that benign diseases are misdiagnosed as malignant tumors, which will lead to wrong diagnosis and wrong surgery. Therefore, there is an urgent need to find a convenient, cost-effective, and noninvasive diagnostic method with very high specificity to substantially reduce the false-positive rate of pancreatic enhancement CT in these special patients or to significantly reduce the false-negative rate while maintaining a high specificity.

Based on the detection of novel tumor markers of miRNA in serum, Shenzhen KeRuiDa Health Technology Co., Ltd. has developed an in vitro diagnostic test, YiDiXie ™ all-cancer test (hereinafter referred to as YiDiXie™ test), which can detect multiple types of cancers with only 200 microliters of whole blood or 100 microliters of serum each time^14^. the YiDiXie ™ test consists of three different tests, YiDiXie™-HS, YiDiXie™-SS and YiDiXie™-D^14^.

The purpose of this study was to evaluate the diagnostic value of YiDiXie™-SS, YiDiXie™-HS, and YiDiXie™-D, in pancreatic cancer.

## PATIENTS AND METHODS

### Study design

This work is part of the sub-study “Evaluating the diagnostic value of the YiDiXie ™ test in multiple tumors” of the SZ-PILOT study (ChiCTR2200066840).

SZ-PILOT is a single-center prospective observational study (ChiCTR2200066840)^14^. Subjects who provided informed agreement to donate their residual samples at the time of admission or during physical examination were considered for inclusion. For this study, a 0.5 milliliter serum sample was collected.

The participants in this investigation were kept blind. the YiDiXie ™ test was conducted by laboratory specialists without knowledge of the subjects’ clinical information. The personnel of KeRuiDa laboratory analyzed the test data. Clinical specialists examining individuals’ clinical data were uninformed of the YiDiXie™ test results.

The study was carried out in accordance with the Declaration of Helsinki and the International Conference on Harmonization (ICH) Code of Practice for the Quality Management of Pharmaceutical Clinical Trials, with approval from the Ethics Committee of Peking University’s Shenzhen Hospital.

### Participants

This study comprised participants with ultrasound-positive pancreatic tumors; the subjects were enrolled individually, and the subjects were continually added as long as they met the inclusion criteria^14^.

Initially included in this study were inpatients with “suspected (solid or blood) malignancy” who signed the informed consent for the remaining samples. Subjects classified as having a malignant tumor (postoperative pathological diagnosis of “malignancy”) or a benign tumor (postoperative pathological diagnosis of “benign tumor”) were assigned to the respective groups. This study did not include the pathology data that were malignant or benign. A portion of the samples from the group studying malignant tumors were utilized in earlier work of our team^14^.

Excluded from this study were subjects who had previously failed the YiDiXie ™ test due to a failure in the serum sample quality test. See our earlier article^14^ for details on enrollment and disqualification.

### Sample collection, processing

There was no need for further blood sampling because the serum samples utilized in this study were from serum that was left over from a typical doctor visit. A volume of 0.5 milliliters was taken out of each person’s leftover serum and kept at -80°C in order to perform the YiDiXie™ test.

### The YiDiXie™ test

The YiDiXie ™ all-cancer detection kit, an in vitro diagnostic kit created and produced by Shenzhen KeRuiDa Health Technology Co., is used in the YiDiXie™ test^14^. In order to identify cancer in participants, this test evaluates the expression levels of several dozen miRNA indicators in serum^14^. Through the integration of separate assays into a contemporaneous testing format and the establishment of appropriate criteria for every miRNA biomarker, the YiDiXie ™ test preserves specificity and improves sensitivity in a broad spectrum of cancer types^14^.

Three separate tests are included in the YiDiXie ™ test: YiDiXie ™ -Super Sensitive (YiDiXie ™ -SS), YiDiXie ™ -Highly Sensitive (YiDiXie ™ -HS), and YiDiXie™-Diagnosis (YiDiXie™-D)^14^. While YiDiXie™ -SS dramatically increases the number of miRNA tests to achieve high sensitivity across all clinical phases of diverse malignant tumor types, YiDiXie™ -HS places a higher priority on specificity and sensitivity^14^.

YiDiXie™-Diagnosis (YiDiXie™-D) ensures high specificity (low misdiagnosis rate) for all cancer types by raising the diagnostic threshold of a single miRNA test^14^.

The YiDiXie ™ all-cancer detection kit’s instructions should be followed when conducting the YiDiXie ™ test^14^. Our previous work^14^ has comprehensive instructions

The laboratory technicians of Shenzhen KeRuiDa Health Technology Co., Ltd evaluated the initial test results once it was finished and classified the YiDiXie ™ test results as “positive” or “negative”^14^.

### Diagnosis of Enhanced CT

The diagnostic conclusion of the enhanced CT examination is judged to be “positive” or “negative”. If the diagnostic conclusion is positive, more positive, or favors malignant tumors, the result is considered “positive”. If the diagnosis is positive, more positive, or favors a benign tumor, or if the diagnosis is ambiguous, the result is considered “negative”.

### Extraction of clinical data

The clinical, pathological, laboratory, and imaging data used in this inquiry came from the individuals’ inpatient medical records or physical examination reports. Clinical staging was conducted by qualified physicians who were evaluated in compliance with the AJCC staging manual (7th or 8th edition)^15-16^.

### Statistical analyses

Descriptive statistics were provided for the demographic variables and baseline attributes. Continuous variables were described by the total number of individuals (n), mean, standard deviation (SD) or standard error (SE), median, first quartile (Q1), third quartile (Q3), minimum, and maximum values. To express categorical variables, the number of individuals in each group and their proportion were utilized. The Wilson scoring method was used to calculate the 95% confidence intervals (CI) for each of the indicators.

## RESULTS

### Participant disposition

62 participants were finally included in this study (malignant group, n=37; benign group, n=25). The demographic and clinical characteristics of the 62 participants in the study are presented in Table 1.

**Table 1.** Participants’s demographic and clinical manifestation.

|  | Malignant (n =37) | Benign (n =25) | Total (N = 62) |
| --- | --- | --- | --- |
| Age, years |  |  |  |
| Mean (SD) | 63.1 (10.95) | 47.1 (17.66) | 56.3 (16.25) |
| Median (Q1,Q3) | 65 (56, 70) | 53 (27, 60) | 58 (52, 66) |
| Min, max | 25, 81 | 21, 76 | 21, 81 |
| Age, group, n (%) |  |  |  |
| < 50 | 2 (5.4) | 12 (48.0) | 14 (22.6) |
| ≥ 50 | 35 (94.6) | 13 (52.0) | 48 (77.4) |
| < 65 | 18 (48.6) | 22 (88.0) | 40 (64.5) |
| ≥ 65 | 19 (51.4) | 3 (12.0) | 22 (35.5) |
| Sex, n (%) |  |  |  |
| Female | 17 (45.9) | 19 (76.0) | 36 (58.1) |
| Male | 20 (54.1) | 6 (24.0) | 26 (41.9) |
| Body mass index (kg/m2) |  |  |  |
| n | 37 | 23 | 60 |
| Mean (SD) | 22.1 (2.32) | 22.1 (3.47) | 22.1 (2.79) |
| Median (Q1,Q3) | 21.5 (20.4, 22.9) | 22.2 (20.6, 24.2) | 21.9 (20.2, 24.0) |
| Min, max | 18.6 28.2 | 15.6 28.5 | 15.6 28.5 |
| Body mass index category, n (%) |  |  |  |
| Underweight | 0 (0) | 4 (16.0) | 4 (6.5) |
| Benign | 28 (75.7) | 12 (48.0) | 40 (64.5) |
| Overweight | 8 (21.6) | 5 (20.0) | 13 (21.0) |
| Obese | 1 (2.7) | 2 (8.0) | 3 (4.8) |
| Missing | 0 (0) | 2 (8.0) | 2 (3.2) |
| AJCC clinical stage |  |  |  |
| Stage I | 11 (29.7) |  | 11 (29.7) |
| Stage II | 9 (24.3) |  | 9 (24.3) |
| Stage III | 6 (16.2) |  | 6 (16.2) |
| Stage IV | 10 (27.0) |  | 10 (27.0) |
| Missing | 1 (2.7) |  | 1 (2.7) |

Q1,Q3, first quartile, third quartile; SD, standard deviation.

In terms of clinical and demographic traits, the two study subject groups were similar (Table 1). The mean (standard deviation) age was 56.3 (16.25) years and 58.1% (36/62) were female.

### Diagnostic performance of YiDiXie™-SS

As shown in Table 2, the sensitivity of YiDiXie™ SS was 100% (95% CI: 90.6% - 100%) and its specificity was 68.0% (95% CI: 48.4% - 82.8%). This means that YiDiXie ™ SS has extremely high sensitivity and high specificity in pancreatic tumors.

**Table 2.**
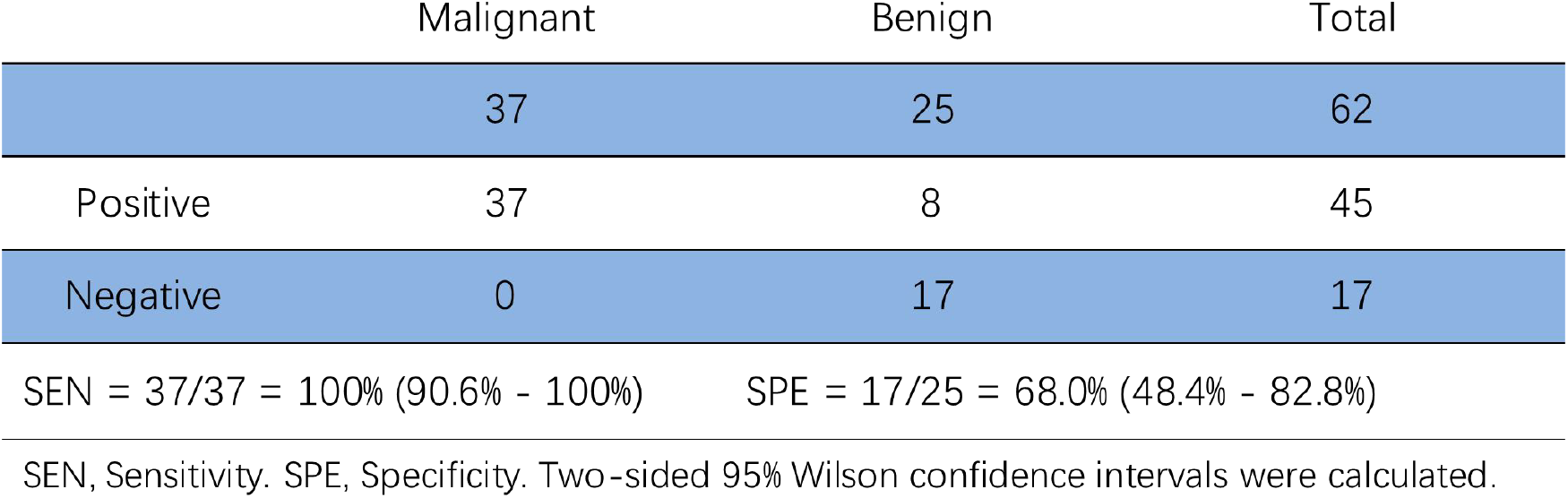
Performance of YiDiXie™-SS.

### Diagnostic performance of YiDiXie™-HS

As shown in Table 3, YiDiXie ™ -HS has a sensitivity of 94.6% (95% CI: 82.3% - 99.0%) and a specificity of 84.0% (95% CI: 65.3% - 93.6%). This means that YiDiXie ™ -HS has high sensitivity and specificity in pancreatic tumors.

**Table 3.**
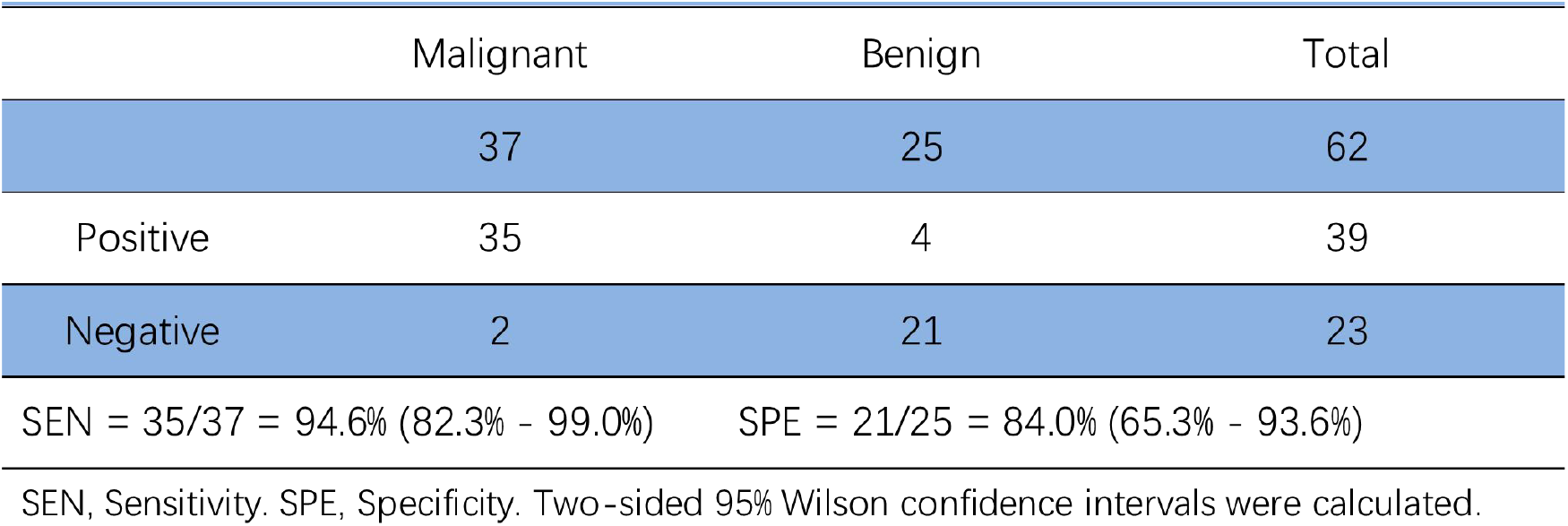
Performance of YiDiXie™-HS.

|  | Malignant | Benign | Total |
| --- | --- | --- | --- |
|  | 37 | 25 | 62 |
| Positive | 35 | 4 | 39 |
| Negative | 2 | 21 | 23 |

SEN = $35/37 = 94.6\%$ (82.3% - 99.0%) SPE = $21/25 = 84.0\%$ (65.3% - 93.6%)
SEN, Sensitivity. SPE, Specificity. Two-sided 95% Wilson confidence intervals were calculated.

### Diagnostic performance of YiDiXie™-D

As shown in Table 4, YiDiXie ™ -D has a sensitivity of 73.0% (95% CI: 57.0% - 84.6%) and its specificity is 92.0% (95% CI: 75.0% - 98.6%). This means that YiDiXie ™ -D has high sensitivity and very high specificity in pancreatic tumors.

**Table 4.** Performance of YiDiXie™-D.

|  | Malignant | Benign | Total |
| --- | --- | --- | --- |
|  | 37 | 25 | 62 |
| Positive | 27 | 2 | 29 |
| Negative | 10 | 23 | 33 |

SEN = 27/37 = 73.0% (57.0% - 84.6%) SPE = 23/25 = 92.0% (75.0% - 98.6%)
SEN, Sensitivity. SPE, Specificity. Two-sided 95% Wilson confidence intervals were calculated.

### Diagnostic performance of enhanced CT

As shown in Table 5, the sensitivity of enhanced CT was 81.1% (95% CI: 65.8% - 90.5%) and its specificity was 76.0%(95% CI: 56.6% - 88.5%).

**Table 5.**
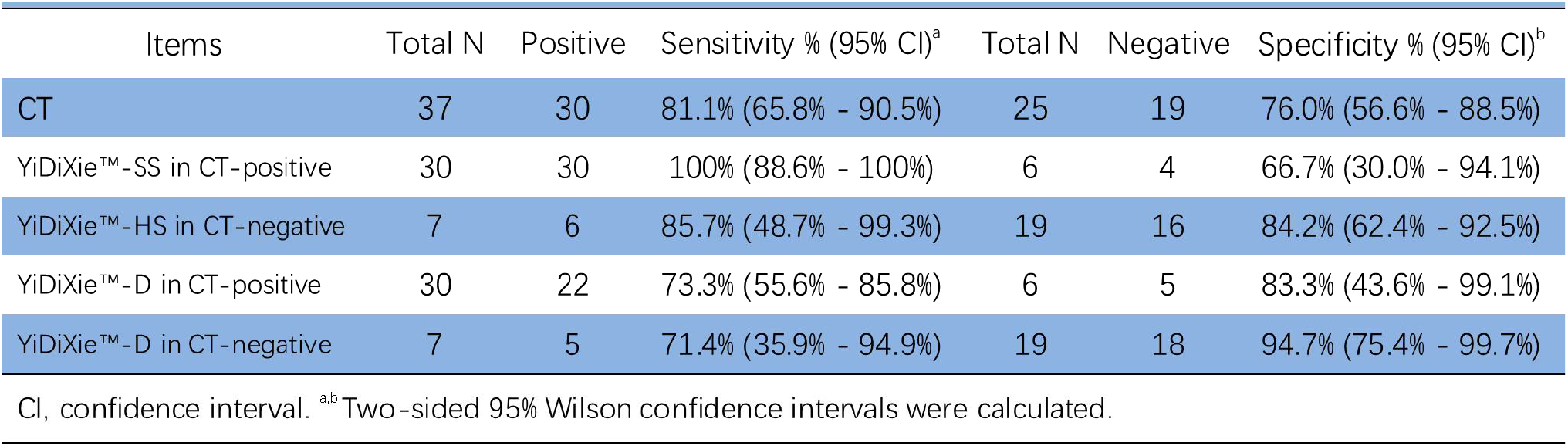
Performance of different items.

| Items | Total N | Positive | Sensitivity % (95% CI)<sup>a</sup> | Total N | Negative | Specificity % (95% CI)<sup>b</sup> |
| --- | --- | --- | --- | --- | --- | --- |
| CT | 37 | 30 | 81.1% (65.8% - 90.5%) | 25 | 19 | 76.0% (56.6% - 88.5%) |
| YiDiXie™-SS in CT-positive | 30 | 30 | 100% (88.6% - 100%) | 6 | 4 | 66.7% (30.0% - 94.1%) |
| YiDiXie™-HS in CT-negative | 7 | 6 | 85.7% (48.7% - 99.3%) | 19 | 16 | 84.2% (62.4% - 92.5%) |
| YiDiXie™-D in CT-positive | 30 | 22 | 73.3% (55.6% - 85.8%) | 6 | 5 | 83.3% (43.6% - 99.1%) |
| YiDiXie™-D in CT-negative | 7 | 5 | 71.4% (35.9% - 94.9%) | 19 | 18 | 94.7% (75.4% - 99.7%) |

CI, confidence interval. <sup>ab</sup> Two-sided 95% Wilson confidence intervals were calculated.

### Diagnostic performance of YiDiXie™-SS in pancreatic enhanced CT-positive patients

In order to solve the challenge of high false-positive rate of pancreas-enhanced CT, YiDiXie ™ -SS was applied to pancreas-enhanced CT-positive patients.

As shown in Table 5, YiDiXie ™ SS had a sensitivity of 100% (95% CI: 88.6% - 100%) and a specificity of 66.7% (95% CI: 30.0% - 94.1%) in patients with positive enhanced CT. This means that the application of YiDiXie ™ SS reduces the false-positive rate of enhanced CT by 66.7% (95% CI: 30.0% - 94.1%) with essentially no increase in malignancy leakage.

### Diagnostic Performance of YiDiXie™-HS in enhanced CT-negative patients

In order to solve the challenge of high rate of missed diagnosis in enhanced CT, YiDiXie™-HS was applied to enhanced CT-negative patients.

As shown in Table 5, the sensitivity of YiDiXie™ -HS was 85.7%(95% CI: 48.7% - 99.3%) and its specificity was 84.2%(95% CI: 62.4% -92.5%).This means that the application of YiDiXie™-HS reduces the false-negative rate of enhanced CT by 84.2%(95% CI: 62.4% -92.5%).

### Diagnostic Performance of YiDiXie™-D in enhanced CT-positive patients

False-positive consequences are significantly worse than false-negative consequences in certain patients with pancreatic tumors, so YiDiXie ™-D is applied to these patients to reduce their false-positive rate.

As shown in Table 5, YiDiXie ™ -D has a sensitivity of 73.3%(95% CI: 55.6% - 85.8%) and a specificity of 83.3%(95% CI: 43.6% - 99.1%) in patients with positive enhancement CT. This means that YiDiXie ™ -D reduces the false positive rate of enhanced CT by 83.3%(95% CI: 43.6% - 99.1%).

### Diagnostic Performance of YiDiXie™-D in enhanced CT-negative patients

Certain enhanced CT-negative patients had significantly worse false-positive than false-negative consequences, so the more specific YiDiXie™-D was applied to such patients.

As shown in Table 5, YiDiXie ™ -D had a sensitivity of 71.4% (95% CI: 35.9% - 94.9%) and a specificity of 94.7% (95% CI: 75.4% - 99.7%) in patients with negative enhanced CT. This means that YiDiXie™-D reduces the false-negative rate of enhanced CT by 71.4% (95% CI: 35.9% - 94.9%) while maintaining high specificity.

## DISCUSSION

### Clinical significance of YiDiXie™-SS in pancreatic enhanced CT-positive patients

The YiDiXie™ test consists of three tests with very different characteristics: YiDiXie™-HS, YiDiXie ™ -SS and YiDiXie ™ -D^14^. YiDiXie ™ -HS balances sensitivity and specificity with high sensitivity and specificity, while YiDiXie ™ -SS has very high sensitivity for all types of malignant tumors, but has a slightly lower level of specificity^14^. The YiDiXie™-D has very high specificity for all malignant tumor types, but low sensitivity^14^.

For patients with positive enhanced CT findings of pancreatic tumors, further diagnostic methods with high sensitivity and specificity are crucial. Balancing these two metrics essentially involves weighing the risks of “missing malignant tumors” against “misdiagnosing benign tumors”. In general, when a benign tumor of the pancreas is misdiagnosed as malignant, it will usually likely lead to unnecessary surgery. However, it will not affect the patient’s prognosis and its treatment is much less expensive than that of advanced cancer. In addition, the positive predictive value is higher in pancreatic enhanced CT-positive patients. Even if the false-negative rate is comparable to the false-positive rate, it can be more harmful. Thus, choosing YiDiXie ™ -SS which offers very high sensitivity but slightly lower specificity, helps reduce the false-positive rate of pancreatic enhanced CT.

As shown in Table 5, YiDiXie ™ -SS had a sensitivity of 100% (95% CI: 88.6% - 100%) and its specificity was 66.7% (95% CI: 30.0% - 94.1%) in patients with positive pancreatic enhanced CT. The above results suggest that YiDiXie™-SS reduces the false-positive rate of pancreas-enhanced CT by 66.7% (95% CI: 30.0% - 94.1%) while maintaining a sensitivity close to 100%.

The above results imply that YiDiXie ™ -SS greatly reduces the probability of incorrectly performing subsequent surgery for benign pancreatic tumors, with essentially no increase in missed diagnosis of malignant tumors. In other words, YiDiXie ™ -SS sharply lowers the mental suffering, expensive examination and surgical costs, surgical injuries, and other adverse consequences for patients with false-positive pancreatic enhanced CT, with essentially no increase in delayed treatment of malignant tumors. Therefore, YiDiXie™-SS well meets the clinical needs and has important clinical significance and wide application prospects.

### Clinical significance of YiDiXie™-HS in enhanced CT-negative patients

For enhanced CT negative patients, further diagnostic methods with high sensitivity and specificity are crucial. Balancing these two metrics inherently involves weighing the risks of “missed malignant tumor diagnoses” against “misdiagnosed benign tumors”. A higher false negative rate implies more missed malignant tumors, leading to delayed treatment, tumor progression, and potentially advanced-stage development. This results in poorer prognosis, shorter survival, reduced quality of life, and increased treatment costs for patients.

In general, when benign pancreatic tumors are misdiagnosed as malignant, radical resection is typically performed without adversely affecting patient prognosis, and treatment costs are far lower than for advanced cancer. Therefore, for CT-negative patients, the risk of “missed malignant tumor diagnoses” outweighs that of “misdiagnosed benign tumors”. Consequently, selecting YiDiXie™-HS, which offers both high sensitivity and specificity, helps reduce false negative rates in pancreatic tumor enhanced CT.

As shown in Table 5, the sensitivity of YiDiXie™ -HS was 85.7%(95% CI: 48.7% - 99.3%) and the specificity was 84.2%(95% CI: 62.4% -92.5%). These results indicate that the application of YiDiXie™-HS reduced the false-negative rate of enhanced CT by 84.2%(95% CI: 62.4% -92.5%).

In summary, YiDiXie™-HS effectively decreases the likelihood of missed malignant tumor diagnoses in CT-negative patients, thereby mitigating adverse outcomes such as poor prognosis, high treatment costs, reduced quality of life, and shorter survival. Therefore, YiDiXie ™ -HS meets clinical needs well, offering significant clinical relevance and broad application prospects.

### Clinical significance of YiDiXie™-D

Pancreatic tumors considered malignant typically undergo curative resection surgery. However, in certain cases, careful consideration is needed before deciding on surgery, such as for smaller tumors or patients in poor general condition.

Further diagnostic methods with high sensitivity and specificity are crucial for these pancreatic tumors patients. Balancing the trade-off between sensitivity and specificity essentially involves weighing the risk of “missing malignant tumors” against “misdiagnosing benign tumors”. Since smaller tumors have a lower risk of tumor progression and distant metastasis, the risk of “missing malignant tumors” is much lower than the risk of “misdiagnosing benign tumors”. For patients in poor general condition, the perioperative risks are significantly higher than for those in general condition, thus the risk of “misdiagnosing benign tumors” is much higher than the risk of “missing malignant tumors”. Therefore, for these patients, YiDiXie ™ -D, which has very high specificity but slightly lower sensitivity, was chosen to reduce the false-positive rate of pancreas-enhanced CT or to significantly reduce its false-negative rate while maintaining high specificity.

As shown in Table 5, the sensitivity of YiDiXie™ -D in patients with positive enhanced CT of the pancreas was 73.3% (95% CI: 55.6% - 85.8%), and its specificity was 83.3% (95% CI: 43.6% - 99.1%); the sensitivity of YiDiXie™-D in patients with negative enhanced CT was 71.4% (95% CI: 35.9% - 94.9%), and its specificity was 94.7% (95% CI: 75.4% - 99.7%). The above results suggest that YiDiXie™-D reduces the false-positive rate of enhanced CT by 83.3% (95% CI: 43.6% - 99.1%) or reduces the false-negative rate of enhanced CT by 71.4% (95% CI: 35.9% - 94.9%) while maintaining a high specificity.

This implies that YiDiXie ™ -D substantially lowers the probability of erroneous surgeries for these patients who require careful consideration. In other words, YiDiXie™-D greatly reduces the risks of surgical trauma, organ removal, pancreatic insufficiency, pancreatic dialysis, and even death, which are serious perioperative complications. Therefore, YiDiXie ™ -D effectively meets clinical needs, offering significant clinical relevance and broad application prospects.

### YiDiXie™ test has the potential to solve two challenges of pancreatic tumor

First of all, the three products of YiDiXie™ test have significant clinical implications in pancreatic tumors. As previously mentioned, YiDiXie™-SS, YiDiXie™-HS and YiDiXie™-D respectively hold important diagnostic value in patients with enhanced CT-positive or negative results.

Second, the three tests of the YiDiXie™ test can significantly alleviate the workload of clinical physicians and facilitate timely diagnosis and treatment of malignancies that were previously subject to delayed treatment. On the one hand, YiDiXie ™ -SS can provide significant relief from non-essential work for surgeons. Patients with Enhanced CT-positive pancreatic tumors usually receive surgical treatment. Whether these surgeries can be completed in a timely manner is directly dependent on the number of surgeons. In many regions of the world, appointments are booked for months or even more than a year. This inevitably delays the treatment of malignant cases among them, and thus it is not unusual for patients with pancreatic tumors awaiting surgery to develop malignant progression or even distant metastases. As shown in Table 5, YiDiXie ™ -SS reduces the false-positive rate of 66.7% (95% CI: 30.0% - 94.1%) in pancreatic enhanced CT-positive patients with essentially no increase in malignant tumor leakage. Thus, YiDiXie ™ -SS can greatly reduce the non-essential workload of surgeons, and facilitate the timely diagnosis and treatment of pancreatic tumors or other diseases that would otherwise be delayed.

On the other hand, YiDiXie™-HS and YiDiXie™-D can significantly alleviate the workload of clinical physicians. In cases where diagnosis with contrast-enhanced CT is difficult, these patients often require contrast-enhanced MRI or pancreatic biopsy. The timely completion of these MRI scans or pancreatic biopsies also depends on the availability of radiologists. In many regions worldwide, appointment delays of several months to over a year are common. Pancreatic tumor patients waiting for contrast-enhanced MRI scans or pancreatic biopsies frequently experience tumor progression to malignancy or distant metastasis. YiDiXie™-HS and YiDiXie™-D can replace these contrast-enhanced MRI scans or pancreatic biopsies, thereby significantly alleviating the workload of clinical physicians and facilitating timely diagnosis and treatment of other malignancies that were otherwise subject to delayed treatment.

Final, the YiDiXie™ test enables “just-in-time” diagnosis of pancreatic tumors. On the one hand, the YiDiXie ™ test requires only microscopic amounts of blood, allowing patients to complete the diagnostic process non-invasively at home. A single YiDiXie™ test requires only 20 microliters of serum, which is equivalent to the volume of 1 drop of whole blood (1 drop of whole blood is about 50 microliters, which produces 20-25 microliters of serum)^14^. Considering the pre-test sample quality assessment experiments and 2-3 repeat experiments, 0.2 ml of whole blood is sufficient to complete the YiDiXie™ test^14^. The 0.2 ml of finger blood can be collected at home using a finger blood collection needle, eliminating the need for venous blood collection by medical personnel and allowing patients to complete the diagnostic process non-invasively without having to leave their homes^14^.

On the other hand, the diagnostic capacity of the YiDiXie™ test is nearly limitless. Figure 1 shows the basic flow chart of the YiDiXie ™ test, which shows that the YiDiXie ™ test requires neither a doctor or medical equipment, nor medical personnel to collect blood^14^. Therefore, the YiDiXie™ test is completely independent of the number of medical personnel and medical facilities, and its testing capacity is nearly unlimited^14^. Thus, the YiDiXie ™ test enables “just-in-time” diagnosis of pancreatic tumors without the patient having to wait anxiously for an appointment.

**Figure 1.**
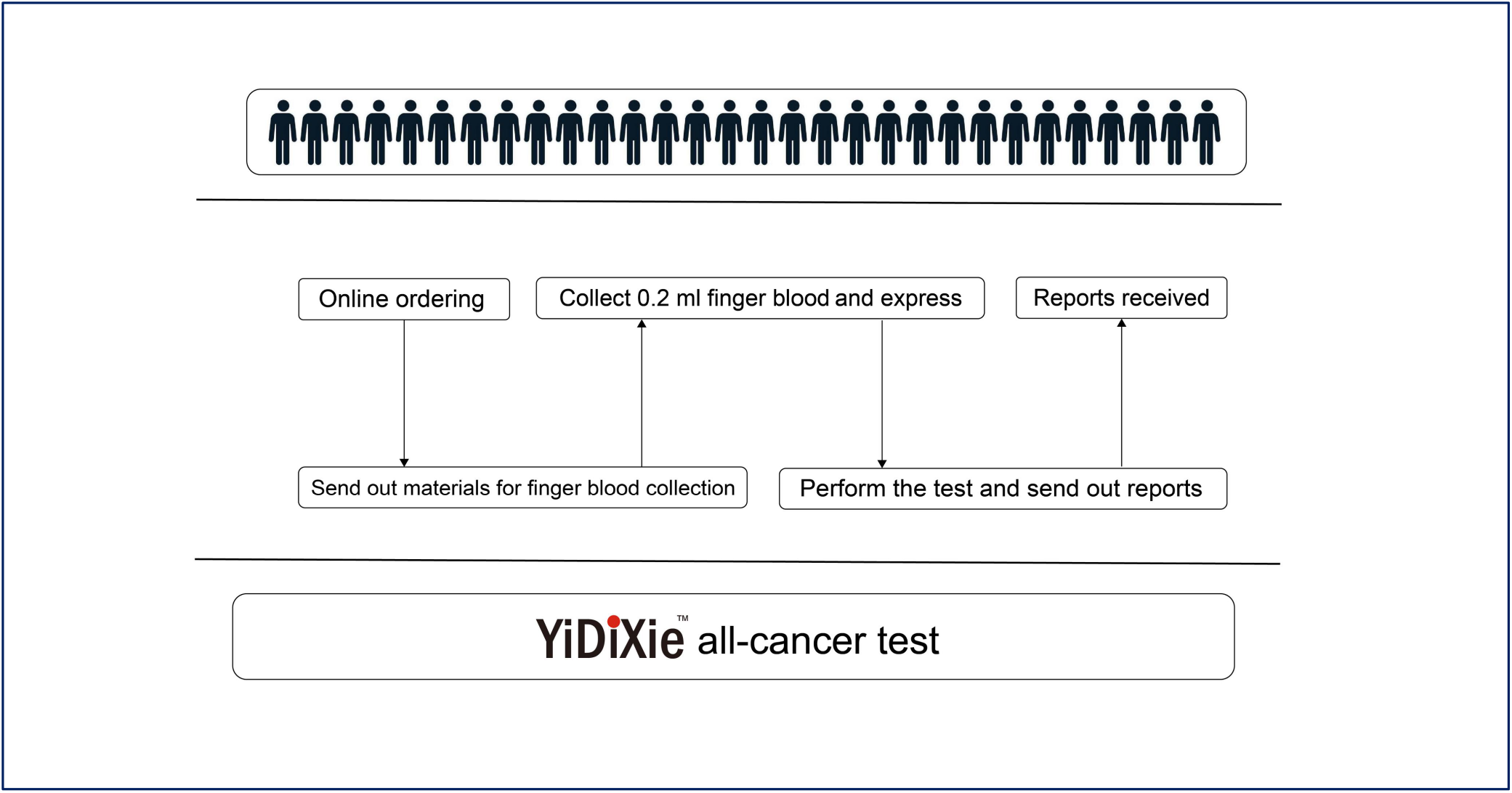
Basic flowchart of the YiDiXie™ test.

In summary, the YiDiXie™ test holds significant diagnostic value for pancreatic tumors, promising to address two challenges, namely, “high false-negative rate of enhanced CT” and “high false-positive rate of enhanced CT”.

### Limitations of the study

First, there were not many patients in this tumor case-benign tumor control study in investigation; therefore, more extensive evaluation will require bigger sample numbers in future clinical trials.

Second, further cohort studies in the natural population of pancreatic tumors are required for further evaluation, as this study was a malignant hospitalized patients.

Last, the fact that this study was conducted in a single center raises the possibility of bias in the findings. Subsequent multi-center research is required for additional assessment.

## CONCLUSION

YiDiXie™-SS has very high sensitivity and high specificity in pancreatic tumors.YiDiXie ™ -HS has high sensitivity and high specificity in pancreatic tumors.YiDiXie ™ -D has high sensitivity and very high specificity in pancreatic tumors.YiDiXie ™ SS significantly reduces pancreatic-enhanced CT with essentially no increase in delayed treatment of malignant tumors. The YiDiXie ™ -HS significantly reduces the false negative rate of pancreatic enhanced CT. the YiDiXie™-D significantly reduces the false positive rate of pancreatic enhanced CT or significantly reduces the false negative rate of pancreatic enhanced CT while maintaining a high level of specificity. the YiDiXie ™ test is of great diagnostic value in pancreatic cancer, and it is expected to solve the problem of “too high a false positive rate” and “too high a false negative rate” of pancreatic enhanced CT.

## Data Availability

All data produced in the present study are contained in the manuscript.

## FUNDING

This study was supported by Shenzhen High-level Hospital Construction Fund, Clinical Research Project of Peking University Shenzhen Hospital (LCYJ2020002, LCYJ2020015, LCYJ2020020, LCYJ2017001).

## Notes

### Competing Interest Statement

The authors have declared no competing interest.

### Clinical Trial

ChiCTR2200066840

### Funding Statement

This work was supported by Shenzhen High-level Hospital Construction Fund, Clinical Research Project of Peking University Shenzhen Hospital (LCYJ2020002, LCYJ2020015, LCYJ2020020, LCYJ2017001).

### Author Declarations

Ethics committee of Peking University Shenzhen Hospital gave ethical approval for this work.

### Summary of Updates

The results were updated. Tabel 2-8 were reviesed.

## REFERENCES

1. Bray F, Laversanne M, Sung H, Ferlay J, Siegel RL, Soerjomataram I and Jemal A: Global cancer statistics 2022: GLOBOCAN estimates of incidence and mortality worldwide for 36 cancers in 185 countries. CA Cancer J Clin. 74: 229–263, 2024.

2. Sung H, Ferlay J, Siegel RL, Laversanne M, Soerjomataram I, Jemal A and Bray F: Global Cancer Statistics 2020: GLOBOCAN Estimates of Incidence and Mortality Worldwide for 36 Cancers in 185 Countries. CA Cancer J Clin. 71: 209–249, 2021.

3. Yamamoto M: A general view of pancreatic cancer in Japan and a proposal for a more practical staging system. International Journal of Clinical Oncology. 4: 267–272, 1999.

4. Yeo CJ and Cameron JL: Improving results of pancreaticoduodenectomy for pancreatic cancer. World J Surg. 23: 907–12, 1999.

5. Alberti A, Dattola A, Parisi A, Maccarone P, Dattola P, Celi S and Basile M: [Periampullar tumors: role of intraoperative color-Doppler ultrasonography in the evaluation of vascular invasion. Methods available to the surgeon and personal experience]. Ann Ital Chir. 71: 669-75; discussion 675-6, 2000.

6. Campen CJ, Dragovich T and Baker AF: Management strategies in pancreatic cancer. Am J Health Syst Pharm. 68: 573–84, 2011.

7. Daamen LA, Groot VP, Goense L, Wessels FJ, Borel Rinkes IH, Intven MPW, van Santvoort HC and Molenaar IQ: The diagnostic performance of CT versus FDG PET-CT for the detection of recurrent pancreatic cancer: a systematic review and meta-analysis. Eur J Radiol. 106: 128–136, 2018.

8. Peti S, Fardanesh R, Golan S, Simpson W, Chin C, Roayaie S, Schwartz M, Labow D and Kostakoglu L: The Combination of FDG PET/CT and Contrast Enhanced CT in the Evaluation of Recurrent Pancreatic Carcinoma and Cholangiocarcinoma. Current Medical Imaging Reviews. 10, 2014.

9. Jung W, Jang JY, Kang MJ, Chang YR, Shin YC, Chang J and Kim SW: The clinical usefulness of 18F-fluorodeoxyglucose positron emission tomography-computed tomography (PET-CT) in follow-up of curatively resected pancreatic cancer patients. HPB (Oxford). 18: 57–64, 2016.

10. Rayamajhi S, Balachandran A, Katz M, Reddy A, Rohren E and Bhosale P: Utility of (18) F-FDG PET/CT and CECT in conjunction with serum CA 19-9 for detecting recurrent pancreatic adenocarcinoma. Abdom Radiol (NY). 43: 505–513, 2018.

11. Seufferlein T, Bachet JB, Van Cutsem E and Rougier P: Pancreatic adenocarcinoma: ESMO-ESDO Clinical Practice Guidelines for diagnosis, treatment and follow-up. Ann Oncol. 23 Suppl 7: vii33–40, 2012.

12. Conroy T, Pfeiffer P, Vilgrain V, Lamarca A, Seufferlein T, O’Reilly EM, Hackert T, Golan T, Prager G, Haustermans K et al.: Pancreatic cancer: ESMO Clinical Practice Guideline for diagnosis, treatment and follow-up. Ann Oncol. 34: 987–1002, 2023.

13. Kitajima K, Murakami K, Yamasaki E, Kaji Y, Shimoda M, Kubota K, Suganuma N and Sugimura K: Performance of integrated FDG-PET/contrast-enhanced CT in the diagnosis of recurrent pancreatic cancer: comparison with integrated FDG-PET/non-contrast-enhanced CT and enhanced CT. Mol Imaging Biol. 12: 452–9, 2010.

14. Chen Sun, Chong Lu, Yongjian Zhang, et al. Evaluation of the Multi-Cancer Early Detection (MCED) value of YiDiXie™-HS and YiDiXie™-SS. medRxiv, 2024:doi: 10.1101/2024.03.11.24303683.

15. Edge SB and Compton CC: The American Joint Committee on Cancer: the 7th Edition of the AJCC Cancer Staging Manual and the Future of TNM. Annals of Surgical Oncology. 17: 1471–1474, 2010.

16. Amin MB, Greene FL, Edge SB, Compton CC, Gershenwald JE, Brookland RK, Meyer L, Gress DM, Byrd DR and Winchester DP: The Eighth Edition AJCC Cancer Staging Manual: Continuing to build a bridge from a population-based to a more “personalized” approach to cancer staging. CA: A Cancer Journal for Clinicians. 67: 93–99, 2017.

